# Effects of virtual fear on real anxiety – A pilot study assessing the effects of exposure in virtual reality and in vivo on anxious healthy participants in narrow rooms

**DOI:** 10.1101/2023.11.20.23298755

**Authors:** Vanessa Renner, Michael Witthöft, Jochen Hardt, Rupert Conrad, Katja Petrowski

**Affiliations:** Medical Psychology, Medical Sociology, University Medical Center of the Johannes Gutenberg-University Mainz, Germany; Department of Clinical Psychology, Psychotherapy, and Experimental Psychopathology, Johannes Gutenberg-University Mainz, Germany; Department of Psychosomatic Medicine and Psychotherapy, University Hospital Muenster, Germany

**Keywords:** anxiety, virtual reality, exposure, heart rate variability

## Abstract

In vivo exposure is a highly effective but rarely implemented treatment for agoraphobia. Most of the patients receive medication or cognitive therapy without exposure because of a high expenditure of money and time for in vivo exposure. Exposure in virtual reality (VR) is easier to implement but the effectiveness of stimulating fear compared to in vivo exposure is still questionable. Therefore, in this study, the effects of in vivo and VR exposure on subjective symptom burden and heart rate variability (HRV) were assessed. 30 healthy individuals with fears in narrow rooms went through in vivo and VR exposure in a randomized order while HRV parameters (RMSSD, HF) and subjective symptom burden was assessed. Linear mixed models were calculated. The effect of condition (VR vs. in vivo), scenario (several narrow rooms) and slot (first 30 seconds, peak, last 30 seconds) on RMSSD and HF was assessed. A random effect for participants (random-intercept term) to allow the intercept to vary across participants was included. Regarding RMSSD and HF, participants showed significantly higher levels during in vivo exposure compared to exposure in VR (RMSSD: p = .005; HF: p < .001), reflecting a stronger activation of the parasympathetic nervous system during in vivo exposure or presumably higher stress levels during VR exposure. This study highlights the necessity of assessing subjective and objective parameters allowing the evaluation of the effectiveness of fear stimulation by exposure approaches. The effectiveness of VR exposure for agoraphobic patients’ needs to be assessed in future studies.

## Introduction

Agoraphobia with or without panic disorder (PD) is characterized by the avoidance of certain places such as busy places, public transport or bridges (Wittchen et al., 2011). Due to the high psychological stress of those affected, they often seek treatment, which makes agoraphobia with or without PD one of the most common diagnoses in outpatient psychotherapy (Wittchen et al., 2011). Cognitive behavioral therapy with exposure to anxiety-inducing situations in vivo (Lang et al., 2009) is considered as “gold standard” of psychotherapeutic treatment of anxiety disorders, as it shows consistently high therapeutic effects (Deacon & Abramowitz, 2004; Ost et al., 2004). However, studies show that 90% of agoraphobic patients receive psychotropic drugs and only 17% are in psychotherapeutic treatment (Margraf & Poldrack, 2000) with only 8% receiving exposure therapy (Becker et al., 2018; Goodwin et al., 2005). Based on a survey, only 13-17% of psychotherapists conduct exposure therapy (Pittig & Hoyer, 2017). Instead, therapists use interoceptive or in sensu exposure and give exposure in vivo as a homework assignment but rarely accompany patients (Hipol & Deacon, 2013; Klan & Hiller, 2014). Due to persistent avoidance behavior and a limited supply of in vivo exposure therapy, the suffering and the likelihood of chronification of the disorder increase (Colman et al., 2007), which in turn creates additional costs for the health care systems (Wittchen et al., 2011).

As Pittig and Hoyer (2017) described, one reason for the low implementation rate of exposure treatments in vivo could be the high expenditure of time and money, which leads to a rapid exhaustion of the therapy quota. This problem could be solved with exposure therapy in virtual reality (VR). A meta-analysis by Eichenberg and Wolters (2012) showed, that exposure therapy in virtual reality (VRET) is effective for the disorders of specific phobia and social anxiety disorder. A reduction in subjectively reported symptoms (e. g. fear, avoidance behavior) and arousal of patients with PD and agoraphobia could be observed after VRET (Botella et al., 2007; Malbos et al., 2013; Pelissolo et al., 2012; Vincelli et al., 2003). In a systematic review by Freitas et al. (2021) regarding the effects of VRET compared with in vivo exposure in anxiety disorders, the authors describe comparable effects of in vivo exposure and VRET in different anxiety disorders. Herein, only two studies with small samples of agoraphobic patients were included (Jang et al., 2000; North et al., 1996). Unfortunately, the results were rarely replicated and only small sample sizes were assessed. However, the major gap in the empirical evidence is the objective proof of similar physiological measured anxiety and arousal between in vivo and VRET.

Emotional processing theory (EPT; Foa & Kozak, 1986) proposes that patients’ fear networks, consisting of propositions related to stimuli and phobic reactions, need to be completely activated during exposure in order to allow therapeutic change. Therefore, it is proposed that physiological activation can serve as an indicator for a successfully activated network and a probable therapeutic change. Therefore, psychophysiological activation during exposure represents an important predictor of therapeutic change (Foa & Kozak, 1986). These assumptions highlight the importance of assessing psychophysiological parameters during exposure.

Heart rate variability (HRV) is an important indicator for measuring stress and anxiety. HRV was already assessed in studies investigating effects of in vivo exposure therapy in anxiety disorders (e. g. Alpers & Sell, 2008; Busscher et al., 2013). In past studies, during exposure, more intense HRV responses to symptom provocation at pre-treatment correlate with a greater symptom reduction in various anxiety disorders (Foa & Kozak, 1986). Some studies found a predictive effect of HR on therapy outcome (Michelson et al., 1990). Though some assumptions of the EPT were not supported in past studies, resulting in the inhibitory learning theory (Craske et al., 2008; Rupp et al., 2017), the assessment of psychophysiological parameters remains relevant to get a holistic picture regarding effects of VR and in vivo exposure.

Therefore, in this study, a sample of healthy participants with fear in narrow rooms as one common fear in agoraphobia was assessed. Effects of VR and in vivo exposure on subjective symptom burden and HRV were compared.

## Method

### Sample

Sample size calculations were conducted using Stata 17.0. A power of 80 %, α = .05, seven points of measurement (baseline plus rooms) and two conditions (VR vs. in vivo) were assumed. Matrices including expected effects per time point and condition were used for calculations. Based on the nature of the scenarios time effects were assumed with no differences between conditions (Carl et al., 2019; Wechsler et al., 2019). Based on this calculation, a sample size of N = 30 resulted. Participants were recruited via announcements at the Johannes Gutenberg-University Mainz, medical practices and online posts. Inclusion criteria were age between 18 and 65 years, ability to speak, read, write in German and giving informed consent. Exclusion criteria were former or current mental disorder, cardiovascular diseases or other diseases influencing the cardiovascular system as well as intake of medication influencing the cardiovascular system. Interested participants were screened for former or current disorders with the SCID-I screening of the DSM-IV (First & Gibbon, 2004; Wittchen et al., 1997). Therein, none of the participants reported to suffer from any past or current mental disorder. Additionally, interested participants were screened for anxiety in narrow rooms based on the Claustrophobia Questionnaire (CLQ; Radomsky et al., 2001). A total of N = 30 participants (86.7 % female) were assessed in this study. Age ranged from 19 to 60 years with M = 30.50 (SD = 12.43). All of the participants provided written informed consent and the study procedure was conducted in accordance with the Declaration of Helsinki and ethically approved by the Landesärztekammer Rheinland-Pfalz, Germany (2020- 15411).

### Procedure

Participants were invited for one appointment lasting two to three hours wherein both, VR and in vivo exposure was conducted in randomized order. First of all, participants were introduced to the nature of fear, explanatory model of anxiety disorders and the idea of exposure. Before starting exposure sessions, participants answered a set of questionnaires (see Clincial assessment) and a three-minute resonance frequency breathing was implemented with a 5 x 5 interval (5 seconds inhale, 5 seconds exhale) was conducted for the baseline HRV assessment. An exposure protocol assessing expected reactions and fears was filled out. Then, the first exposure session started either in VR or in vivo depending on randomization. Afterwards, an exposure protocol assessing actual experiences during exposure was filled out before starting with the second exposure followed by answering the questionnaires and exposure protocols a second time.

In VR, six different real-life scenarios of a narrow basement room were presented after each other. Scenarios were recorded using a 360° camera in a 2-3m^2^ basement room without windows and with a ladder leading to a hatch as the exit. The scenarios differed regarding possibility to flight (hatch open/ closed and ladder present or not) and light (bright, dimmed, dark). Scenarios were not randomized in their order of presentation. First, participants were in the bright room with accessible ladder and opened hatch and last, the hatch was closed, the ladder absent and the light was dimmed. The room for in vivo exposure was also 2-3m^2^ in the fifth floor without any window. To parallelize the in vivo exposure with VR exposure, participants went through six different stages where the door of the room was open/ ajar/ shut/locked and the light was on/ off. As in VR, in vivo participants started with a lighted room, door opened and ended with a locked door in a dark room.

### Clinical assessment

Before and after each exposure session, the CLQ (Radomsky et al., 2001) was used describing 26 situations were claustrophobic fears can occur. Participants rate these situations from 0 (not at all anxious) to 4 (extremely anxious). Internal consistency for the CLQ is high with α = .95 (Radomsky et al., 2001). Additionally, when entering and leaving a room, subjective anxiety levels were assessed on a scale from 0 to 100%.

After VR exposure sessions, participants answered the Igroup presence questionnaire (IPQ; Schubert, 2003) and the Simulation Sickness Questionnaire (SSQ; Neukum & Grattenthaler, 2006). The IPQ (Schubert, 2003) measures the sense of presence experienced in a virtual environment. It includes three subscales (spatial presence, involvement, experienced realism) which can be combined into an overall score. The IPQ meets good internal consistency with Cronbach’s α = .87 (Schubert, 2003). The SSQ (Neukum & Grattenthaler, 2006) assesses sickness symptoms such as headache, nausea or dizziness elicited by VR. Participants rate the occurrence of 16 different symptoms on a scale from 0 (no perception) to 3 (severe perception). Scores for nausea, oculomotor disturbance, disorientation and total simulator sickness can be computed. Cronbach’s α = .87 (Bouchard et al., 2007).

### Technical devices and Heart Rate Variability

For VR exposure, a standalone Oculus Quest by Meta was used. HRV was obtained with a Polar Watch V800 assessing HRV with a sensor on a chest strap. Intervals for HRV assessment were three minutes and started with entering a new room and ended with leaving it. Additionally, slots were calculated reflecting the first and last 30 seconds in one room to assess possible habituation effects as well as one 30 seconds slot reflecting the peak RMSSD / HF within one room. Peak slots were not calculated for resonance frequency breathing condition. To eliminate extra beats or erroneous values of the R–R interval data, the software Polar ProTrainer 5 (Polar, Germany) was used to post-process the recordings by an automatic filtering process method (filter power: moderate, minimum protection zone: 6 sqm). The root-mean-square successive differences parameter (RMSSD) to reflect the activity of the parasympathetic nervous system as well as the heart frequency (Hf) were calculated. Analyses of the parameters were conducted with Kubios HRV Standard (Tarvainen et al., 2014). RMSSD and HF were transformed using logarithm naturalis.

### Statistical analyses

Linear mixed models (LMM) as recommended when analyzing nested data (Hox et al., 2017) were calculated using Jamovi 2.3 (The Jamovi Project, 2020). The effect of condition (VR vs. in vivo), rooms and slot (first 30 seconds, peak, last 30 seconds) on RMSSD and HF serving as dependent variables was assessed. Here, RMSSD and HF repeated measurement points were nested in participants as level 2 variables. A random effect for participants (random-intercept term) to allow the intercept to vary across participants was included. Three models were calculated, one for each HRV parameter. Non-significant interactions within the models were excluded stepwise, highest p-values were excluded first.

## Results

All of the participants showed values above the established cut-off for a high risk for claustrophobic events in narrow rooms of M = 0.33 in the CLQ (Napp et al., 2017, 2021) before exposure sessions ranging from M = 0.53 to M = 1.24. Figure 1 shows subjective levels of anxiety for each room during VR and in vivo exposure. Figure 2 and 3 show percentages of subjective anxiety when entering and leaving the rooms during both exposure sessions.

**Figure 1.**
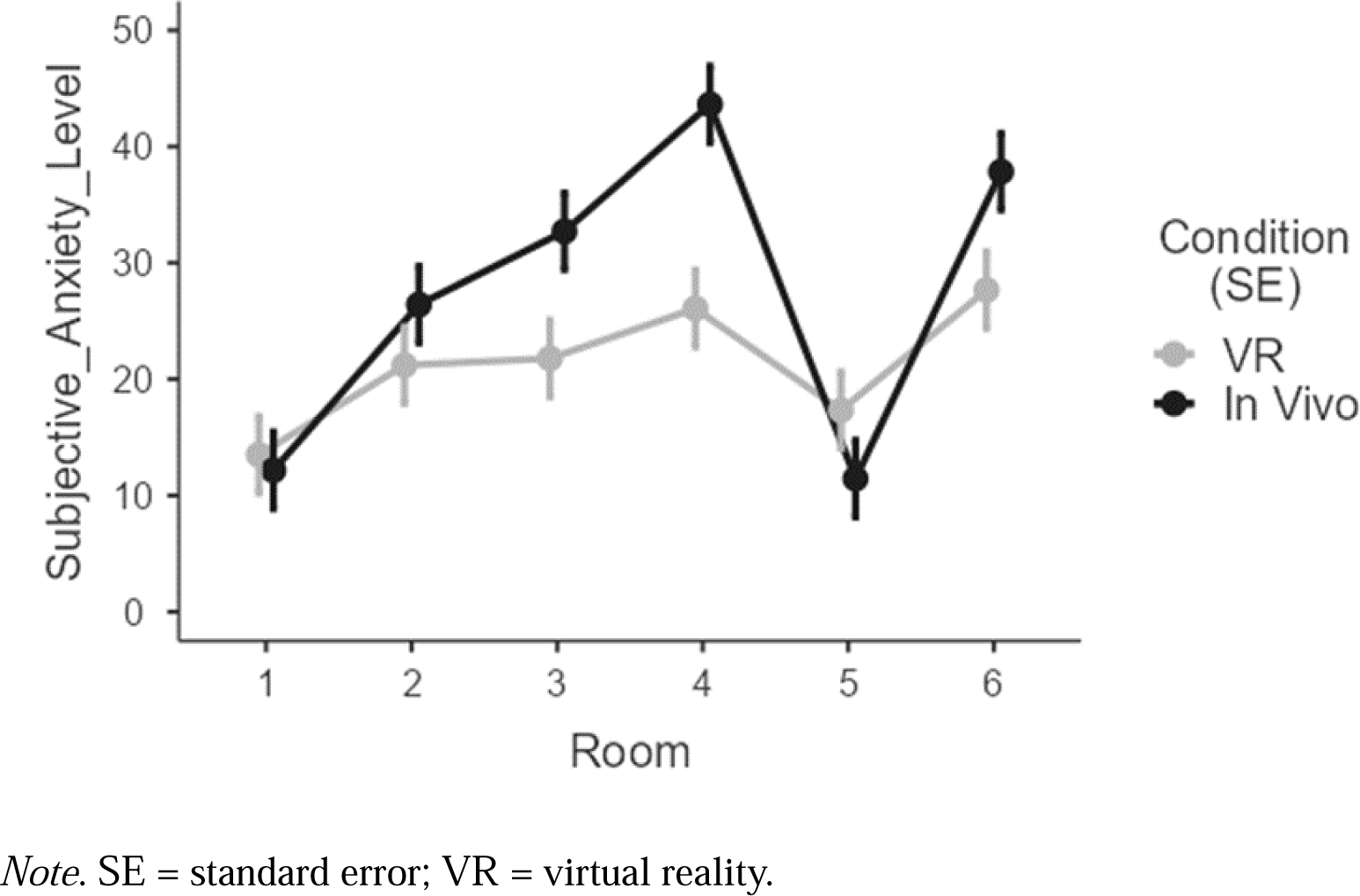
Percentages of subjective anxiety ratings for each room.

**Figure 2.**
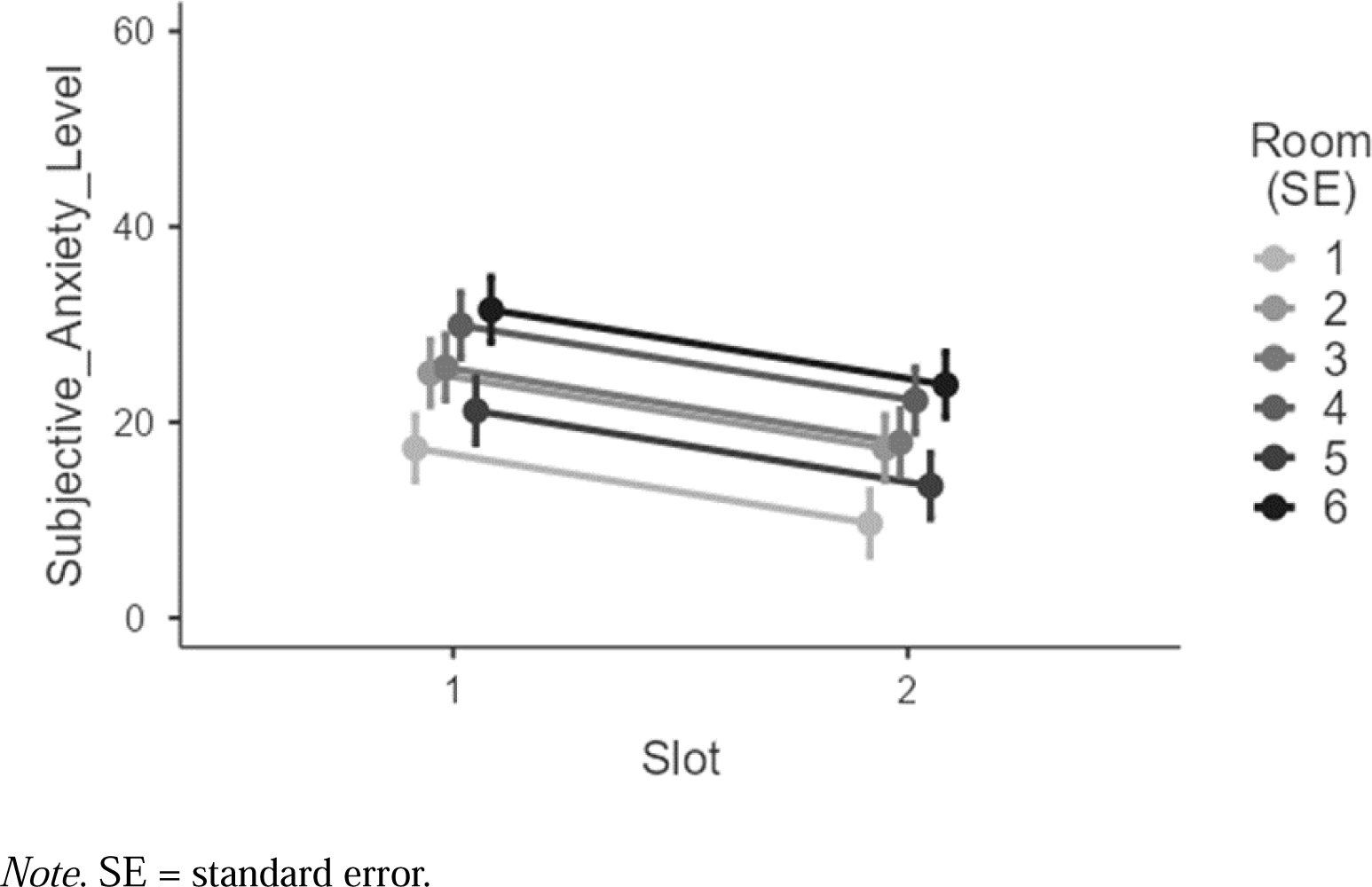
Percentages of subjective anxiety ratings when entering and leaving each room during VR exposure.

**Figure 3.**
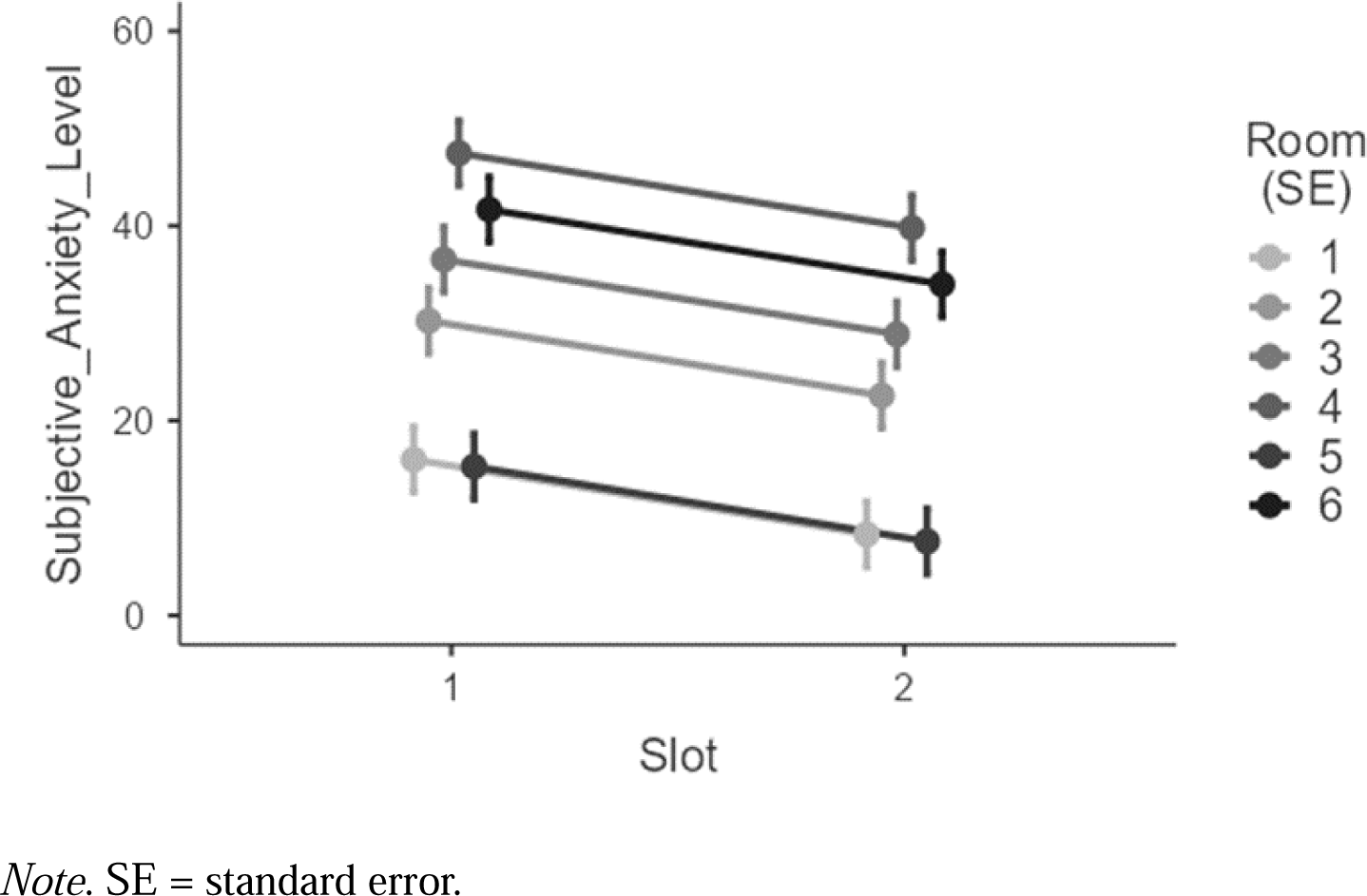
Percentages of subjective anxiety ratings when entering and leaving each room during in vivo exposure.

After VR exposure, the mean IPQ score was M = 2.98 (SD = 0.65) ranging from M = 1.69 to M = 4.15. A total of 53.3% did report a general sense of presence in the VR environment of at least M = 3.00. The mean score of spatial presence was M = 2.84 (SD = 0.50), of involvement M = 3.16 (SD = 0.57) and experienced realism M = 2.84 (SD = 1.33). Regarding the SSQ, the most problems occurred regarding oculomotor disturbances (M = 4.96, SD = 3.39) followed by disorientation (M = 3.57, SD = 3.47) and nausea (M = 3.33, SD = 4.15). In total, 6.7% did not report any symptoms of simulation sickness, 73.3% of the participants scored smaller than 10. The maximum score on the SSQ was 27 out of 48.

Mean scores of RMSSD and HF for resonance frequency breathing and exposure rooms can be derived from table 1 and 2. Regarding RMSSD, there is a significant main effect of condition (F(1, 1161.00) = 7.96, p = .005) with higher RMSSD during in vivo exposure compared to VR exposure (see figure 4). There is a significant main effect for slot (F(2, 1161.00) = 18.94, p < .001) due to the first 30 seconds in a room significantly differing from peak RMSSD (t(1161.00) = -5.75, p < .001). There is no significant difference between the first and last 30 seconds in one room (t(1161.00) = -0.97, p = .333) and no main effect of room was found (F(6, 1161.00 = 0.42, p = .863). Fixed effects parameter estimations for RMSSD can be derived from table 2.

**Figure 4.**
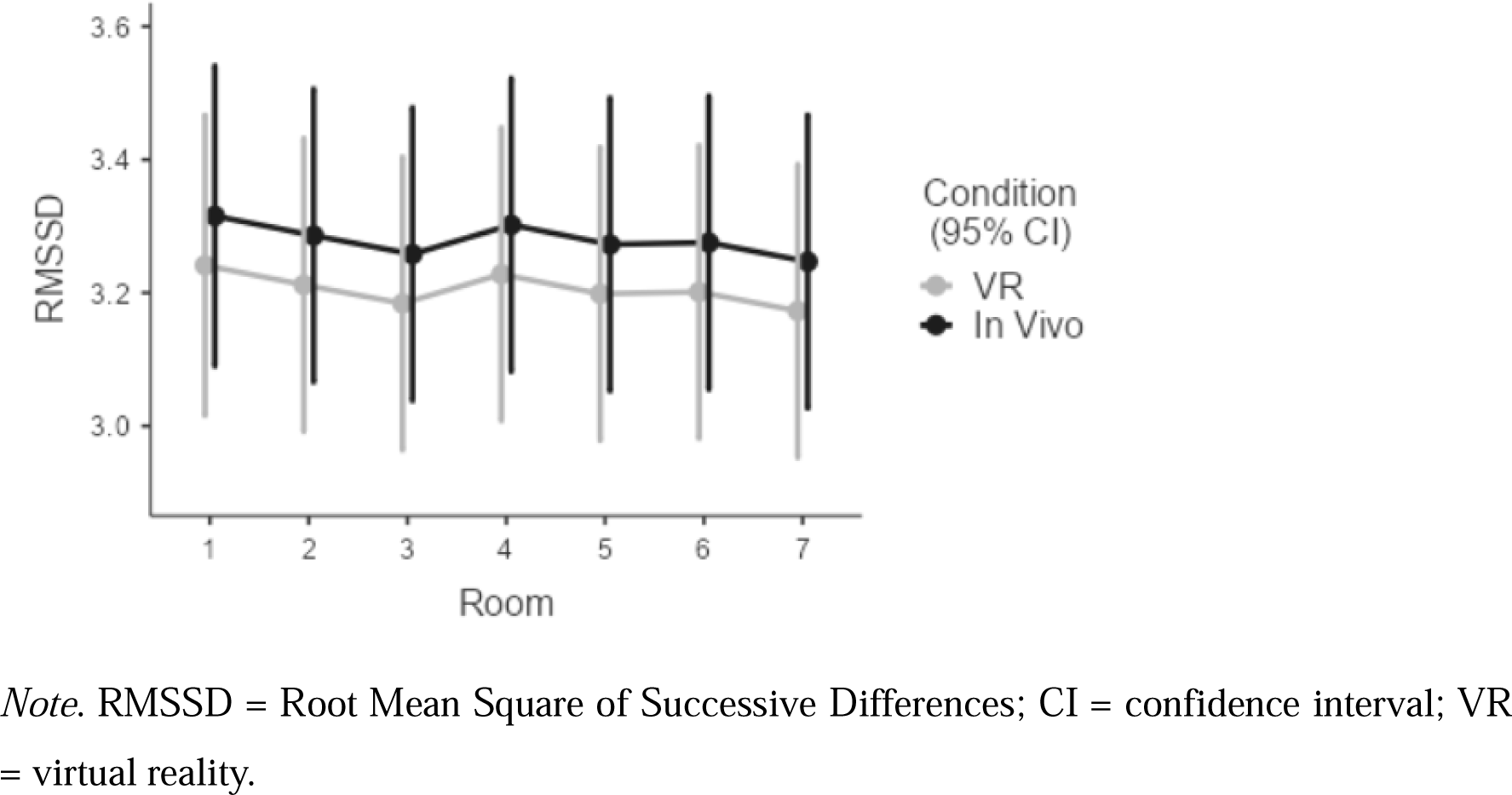
RMSSD for each room during VR and in vivo exposure.

**Table 1.** Descriptives RMSSD.

|  | **N** | **Mean** | **SD** | **Minimum** | **Maximum** |
| --- | --- | --- | --- | --- | --- |
| **VR** |  |  |  |  |  |
| RFB | 30 | 39.69 | 27.74 | 4.83 | 145.29 |
| Room 1 | 30 | 33.72 | 26.96 | 3.69 | 153.88 |
| Room 2 | 30 | 31.16 | 20.54 | 5.24 | 111.93 |
| Room 3 | 30 | 33.02 | 22.45 | 6.12 | 127.49 |
| Room 4 | 30 | 33.45 | 21.67 | 8.50 | 122.43 |
| Room 5 | 30 | 34.02 | 26.94 | 4.98 | 149.38 |
| Room 6 | 30 | 34.87 | 26.25 | 5.38 | 146.90 |
| **In Vivo** |  |  |  |  |  |
| RFB | 30 | 39.67 | 23.24 | 5.17 | 111.75 |
| Room 1 | 30 | 36.61 | 22.72 | 6.43 | 111.55 |
| Room 2 | 30 | 35.38 | 22.55 | 6.50 | 111.24 |
| Room 3 | 30 | 36.02 | 20.06 | 6.82 | 90.24 |
| Room 4 | 30 | 37.92 | 30.39 | 4.13 | 161.76 |
| Room 5 | 30 | 31.50 | 16.79 | 6.06 | 74.58 |
| Room 6 | 30 | 33.53 | 17.92 | 13.70 | 85.88 |

Note. RFB = resonance frequency breathing; RMSSD = Root Mean Square of Successive Differences; VR = virtual reality; SD = standard deviation.

**Table 2.** Descriptives HF.

|  | **N** | **Mean** | **SD** | **Minimum** | **Maximum** |
| --- | --- | --- | --- | --- | --- |
| **VR** |  |  |  |  |  |
| RFB | 30 | 631.17 | 725.77 | 8.25 | 2893.48 |
| Room 1 | 30 | 458.94 | 494.83 | 2.03 | 1800.27 |
| Room 2 | 30 | 401.73 | 387.10 | 5.00 | 1733.35 |
| Room 3 | 30 | 505.20 | 581.71 | 5.91 | 2528.67 |
| Room 4 | 30 | 490.01 | 568.76 | 32.32 | 2545.21 |
| Room 5 | 30 | 417.01 | 457.63 | 8.64 | 1739.12 |
| Room 6 | 30 | 538.08 | 580.20 | 5.15 | 2343.35 |
| **In Vivo** |  |  |  |  |  |
| RFB | 30 | 783.19 | 870.27 | 16.88 | 4334.22 |
| Room 1 | 30 | 599.46 | 642.08 | 7.88 | 2435.23 |
| Room 2 | 30 | 594.64 | 625.97 | 2.47 | 2699.80 |
| Room 3 | 30 | 553.37 | 588.27 | 20.26 | 2191.17 |
| Room 4 | 30 | 950.13 | 2478.57 | 0.44 | 13802.97 |
| Room 5 | 30 | 459.76 | 474.82 | 10.13 | 1821.66 |
| Room 6 | 30 | 536.98 | 738.45 | 37.30 | 3663.99 |

Note. RFB = resonance frequency breathing; HF = heart frequency; VR = virtual reality; SD = standard deviation.

**Table 3.** Fixed Effects Parameter Estimates for RMSSD.

| Names | Effect | Estimate | SE | 95% Confidence Interval |  | df | t | p |
| --- | --- | --- | --- | --- | --- | --- | --- | --- |
|  |  |  |  | Lower | Upper |  |  |  |
| (Intercept) | (Intercept) | 3.24 | 0.10 | 3.04 | 3.44 | 29.03 | 31.53 | <□.001 |
| Condition | In Vivo - VR | 0.07 | 0.03 | 0.02 | 0.13 | 1161.00 | 2.82 | 0.005 |
| Room | 2 - 1 | -0.03 | 0.05 | -0.14 | 0.08 | 1161.00 | -0.54 | 0.589 |
|  | 3 - 1 | -0.06 | 0.05 | -0.16 | 0.05 | 1161.00 | -1.05 | 0.293 |
|  | 4 - 1 | -0.01 | 0.05 | -0.12 | 0.09 | 1161.00 | -0.25 | 0.806 |
|  | 5 - 1 | -0.04 | 0.05 | -0.15 | 0.06 | 1161.00 | -0.78 | 0.433 |
|  | 6 - 1 | -0.04 | 0.05 | -0.15 | 0.07 | 1161.00 | -0.73 | 0.463 |
|  | 7 - 1 | -0.07 | 0.05 | -0.18 | 0.04 | 1161.00 | -1.26 | 0.209 |
| Slot | 2 - 1 | -0.19 | 0.03 | -0.26 | -0.13 | 1161.00 | -5.75 | <□.001 |
|  | 3 - 1 | -0.03 | 0.03 | -0.09 | 0.03 | 1161.00 | -0.97 | 0.333 |

Note. RMSSD = Root Mean Square of Successive Differences; VR = virtual reality; SE = standard estimation; df = degrees of freedom.

Regarding HF, there is a significant main effect for condition (F(1, 1161.00) = 17.73, p < .001) and slot (F(2, 1161.00) = 31.08, p < .001) but not for room (F(6, 1161.00) = 1.45, p = .191). During in vivo exposure, participants showed significantly higher HF than during VR exposure (see figure 5). The first 30 seconds in a room significantly differed from peak HF (t(1161.00) = 7.71, p < .001) and peak significantly differed from the last 30 seconds in the same room (t(1161.00) = -5.67, p < .001). Fixed effects parameter estimations for HF can be derived from table 4.

**Figure 5.**
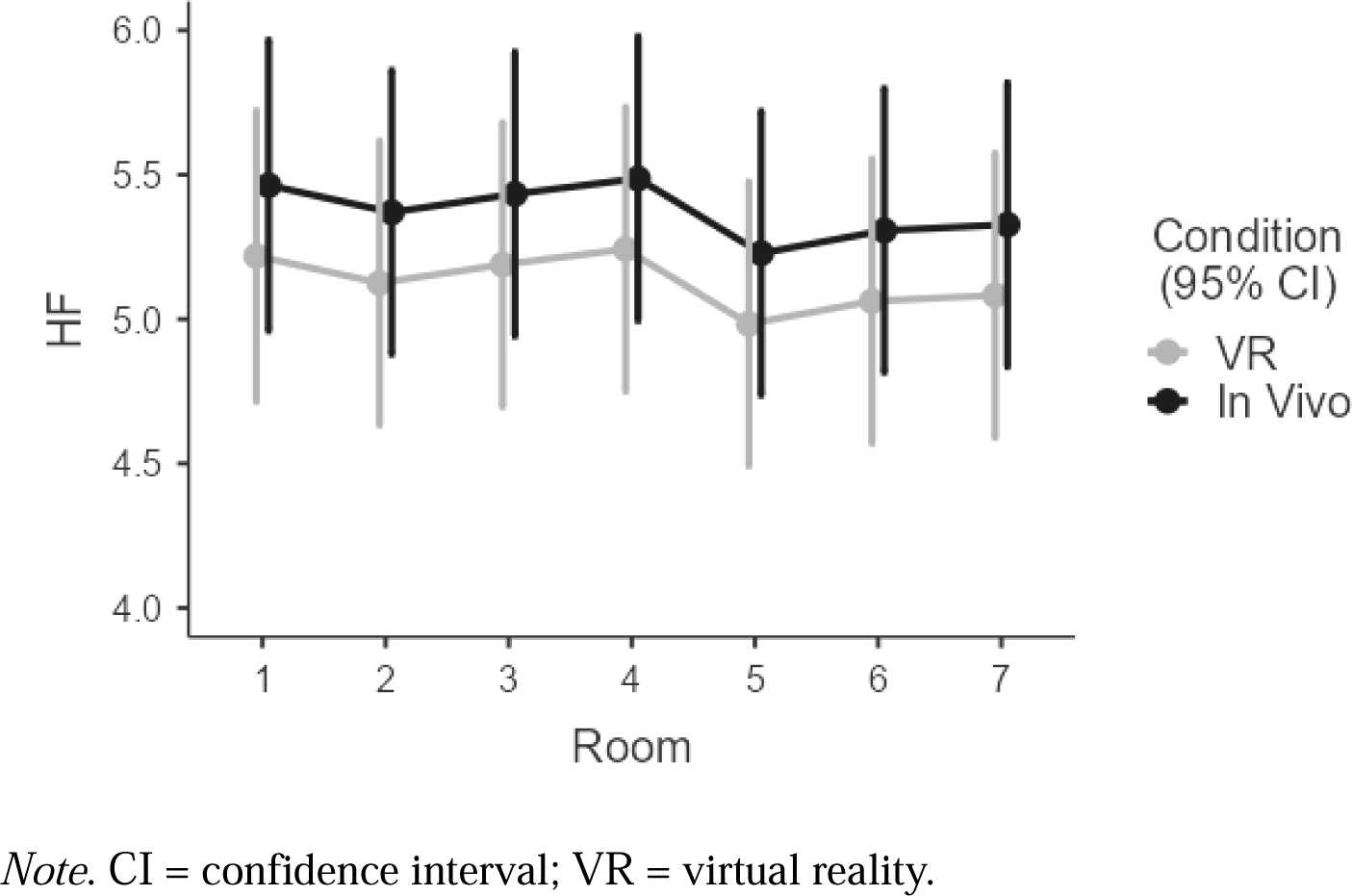
Heart frequency for each room during VR and in vivo exposure.

**Table 4.** Fixed Effects Parameter Estimates for Heart Frequency.

| Names | Effect | Estimate | SE | 95% Confidence Interval |  | df | t | p |
| --- | --- | --- | --- | --- | --- | --- | --- | --- |
|  |  |  |  | Lower | Upper |  |  |  |
| (Intercept) | (Intercept) | 5.25 | 0.23 | 4.80 | 5.70 | 29.03 | 22.83 | <□.001 |
| Condition | In Vivo - VR | 0.24 | 0.06 | 0.13 | 0.36 | 1161.00 | 4.21 | <□.001 |
| Room | 2 - 1 | -0.09 | 0.12 | -0.33 | 0.14 | 1161.00 | -0.77 | 0.439 |
|  | 3 - 1 | -0.03 | 0.12 | -0.27 | 0.21 | 1161.00 | -0.25 | 0.805 |
|  | 4 - 1 | 0.02 | 0.12 | -0.21 | 0.26 | 1161.00 | 0.20 | 0.844 |
|  | 5 - 1 | -0.23 | 0.12 | -0.47 | 0.00 | 1161.00 | -1.95 | 0.052 |

**Table 4.**
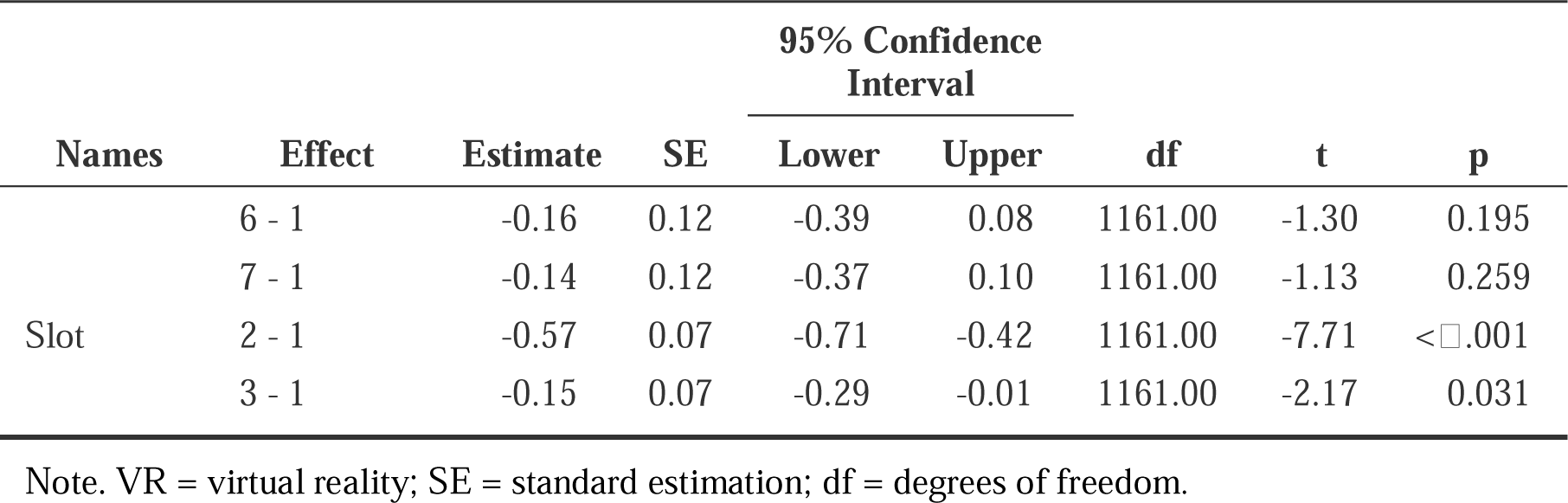
Fixed Effects Parameter Estimates for Heart Frequency.

## Discussion

In this highly standardized and controlled study, the effects of exposure sessions in VR and in vivo were compared regarding subjective symptom burden as well as HRV parameters. The induced fear significantly differed for the different intensities of the stimuli: fully lighted rooms with options to escape (rooms 1, 2, 3) induced less fear than rooms with dimmed light and no escape options induced higher ratings of fear (room 4, 5). An almost completely dark room (room 6) on the other hand, did induce low levels of fear probably because participants did hardly see the narrow room and possible escape options. In general, rooms in vivo did lead to higher anxiety ratings than rooms in VR, except for room 1 and 5. Though participants rated in vivo rooms as more fear-inducing, VR led to more stress reflected by lower levels of RMSSD and HF. This could be due to differences in cognitive processing of VR and in vivo exposure whereby participants expect VR scenarios to be less frightening than real-life situations and therefore give lower ratings.

Regarding HRV parameters (RMSSD, HF), participants showed significantly higher levels during in vivo exposure compared to VR exposure reflecting a higher parasympathetic activation and a more relaxed status of the individual during in vivo exposure than during VR exposure. Though participants rated rooms different regarding fear-inducing potential, this is not shown in HRV parameters where there were no significant differences between rooms. Accordingly, subjective and objective measures of anxiety and ANS-activation differ. In general, it was shown that in subjective ratings, in vivo exposure sessions were more fear-inducing than VR scenarios but during in vivo exposure, not only the physiological arousal but also the parasympathetic activation was higher than during VR.

Based on EPT (Foa & Kozak, 1986), an activation of the autonomous nervous system is necessary for habituation and a positive therapy outcome. In this study, parameters reflecting parasympathetic activation were higher during in vivo exposure but reflect a more relaxed and less activated/stressed status of the individuals during exposure. As exposure sessions are supposed to, at least at first, stress individuals to reach a habituation this finding rather supports VR exposure showing a reduced activation of the parasympathetic nervous system. HRV was also shown to be a biomarker for emotion regulation whereby greater HRV was associated with better top-down self-regulation such as emotion regulation (Holzman & Bridgett, 2017). Therefore, during in vivo exposure, participants could have shown stronger emotion regulation processes to cope with this situation.

The major strength of this study is the highly standardized and controlled setting. Furthermore, the different degrees in intensity of the stimuli made it possible to observe and intensity progress. Unfortunately, the present study shows some limitations. First of all, ratings of subjective anxiety during exposure where rather low. It is assumed, that this is mainly because of the healthy sample not showing any diagnosis of anxiety disorder but elevated levels of fear in narrow rooms.

In future studies, findings of this study should be addressed by 1) assessing patients with agoraphobia regarding subjective symptom burden and objective HRV parameters during exposure sessions, 2) implementing VR and in vivo sessions over a longer period of time (e. g. exposure therapy) and 3) addressing underlying mechanisms promoting effective exposure based on EPT and inhibitory learning.

## Conclusions

In this study, participants showed a higher parasympathetic activation during in vivo exposure than during VR exposure and subjective anxiety ratings were higher for in vivo exposure compared to VR. This finding implies the necessity to further evaluate the effects of VR exposure compared to in vivo exposure and the underlying mechanisms favoring a positive therapy outcome. The present study gives first insights in the potential of VR exposure as add-on or alternative to in vivo exposure showing mixed results and pointing at challenges and future directions for the implementation of VR exposure.

## Data Availability

All data produced in the present study are available upon reasonable request to the authors

## Acknowledgements

We express our gratitude to Eva-Maria Schneider and Florian Mies, both medical doctors, for their unwavering dedication and invaluable support in collecting and processing data. We are also grateful to all the participants who willingly participated in this study, contributing to its success.

